# CircWaveNet: A New Conventional Neural Network Based on Combination of Circlets and Wavelets for Macular OCT Classification

**DOI:** 10.1101/2023.09.23.23295997

**Authors:** Roya Arian, Alireza Vard, Rahele Kafieh, Gerlind Plonka, Hossein Rabbani

## Abstract

Computer-aided systems can help the ophthalmologists in early detection of most of ocular abnormalities using retinal OCT images. The need for more accurate diagnosis increases the need for modifications and innovations to current algorithms. In this paper, we investigate the effect of different X-lets on the classification of OCT B-scans of a dataset with one normal class and two abnormal classes. Different transforms of each B-scan have been fed to the designed 2D-Convolutional-Neural-Network (2D-CNN) to extract the best-suited features. We compare the performance of them with MSVM and MLP classifiers. Comparison with the accuracy of normal and abnormal classes reveals substantially better results for normal cases using 2D-Discrete-Wavelet-Transform (2D-DWT), since the structure of most normal B-scans follows a pattern with zero-degree lines, while for abnormalities with circles appearing in the retinal structure (due to the accumulation of fluid), the circlet transform performs much better. Therefore, we combine these two X-lets and propose a new transform named CircWave which uses all sub-bands of both transformations in the form of a multi-channel-matrix, with the aim to increase the classification accuracy of normal and abnormal cases, simultaneously. We show that the classification results obtained based on CircWave transform outperform those based on the original images and each individual transform. Furthermore, the Grad-CAM class activation visualization for B-scans reconstructed from half of the CircWave sub-bands indicates a greater focus on appearing circles in abnormal cases and straight lines in normal cases at the same time, while for original B-scans the focus of the heat-map is on some irrelevant regions. To investigate the generalizability of our proposed method we have applied it also to another dataset. Using the CircWave transform, we have obtained an accuracy of 94.5% and 90% for the first and second dataset, respectively, while these values were 88% and 83% using the original images. The proposed CNN based on CircWave provides not only superior evaluation parameter values but also better interpretable results with more focus on features that are important for ophthalmologists.

## 1. Introduction

OCT is an imaging technique to provide information about the cross-sectional structure of tissues. This non-invasive method has been widely used in ophthalmology and the investigation of retinal diseases and glaucoma due to the layered structure of the retina(1). OCT is similar to ultrasound imaging technique, however it uses near-infrared light instead of sound beams(2).

Identifying early symptoms of macular degeneration that affect central vision can prevent vision loss, and OCT images can play an important role in this identification since they can demonstrate structural changes in the retina. The most common retinal diseases are age-related macular degeneration (AMD) and diabetic macular edema (DME) (3, 4). AMD is a visual disorder caused by retinal abnormalities that reduces the central vision (5). DME is another retinal disease that is associated with diabetic retinopathy and a leading cause of vision loss for people with diabetes. In this abnormality excess blood glucose damages blood vessels in the retina, causing them to leak. This leakage causes an accumulation of fluid in the macula that makes it swell (4, 6). Figure 1 shows an example of normal, DME, and AMD B-scans.

**Figure 1.**
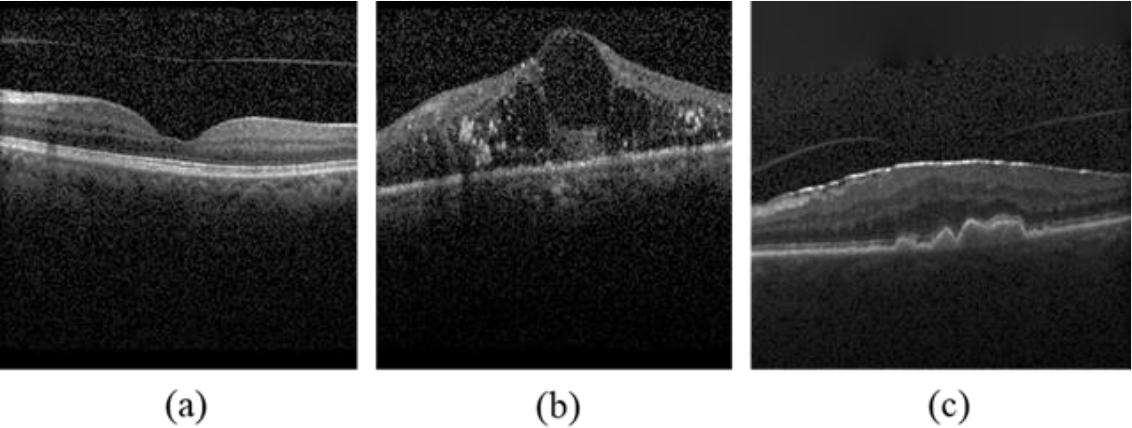
An example of (a) normal, (b) DME and (c) AMD B-scans

Therefore, OCT can be regarded as a remarkable biomarker for the quantitation of AMD and DME disorders. Spectral Domain OCT (SD-OCT) and Swept-Source OCT (SS-OCT) are two newer generations of OCT that offer some advantages including a better rate of acquisition and resolution, less light scattering, providing more clear retinal structural information, and improved speed(7, 8). Notwithstanding these recent advances in OCT technology, still, manual analysis remains time-consuming and error-prone due to the similarity of different abnormalities in OCT images. To address these problems, artificial intelligence algorithms including machine learning and deep learning have been widely used in image processing for different applications such as classification, segmentation, denoising, and compressive sensing(9, 10).

Models used in medical image processing are often based on suitable transforms which are able to exploit correlations of the image data and can therefore lead to sparse image representations (11). In other words, the significant information on the data can be stored already by a small amount of coefficients in the transform domain. Then the classification of this small number of coefficients is much easier and faster.

Depending on the characteristics of the data and the intended application, the appropriate transform needs to be selected.

As mentioned in (11), transform domain approaches can be grouped into data-adaptive and non-data-adaptive models. For classification, non-data-adaptive transforms have the advantage that the information in the transform domain stays to be comparable. Among non-data-adaptive models, X-let transforms based on multi-scale time/space-frequency analysis are very powerful since these transforms connect frequency and time information.

X-lets have been recently used for various OCT image processing applications. Recent approaches that combine deep learning and X-let transforms are summarized in Table 1.

**Table. 1.** Recent researches on using combination of deep learning and different X-lets for image processing.

| Author | year | Application | Dataset | X-let | Combination method | X-let Advantages |
| --- | --- | --- | --- | --- | --- | --- |
| Nur et, al.(12) | 2023 | Classification | CT | Contourlet | Application of Contourlet transform and CNN to extract features individually from segmented images and combining them in one feature vector. | Overcoming the limitations of the tensor-product discrete wavelet transform (DWT) by trying to capture curves rather than points singularities |
| Tian et, al.(13) | 2023 | Denosing | Natural images (BSD) | Wavelet | Multi-stage image denoising CNN with the wavelet transform via three stages, i.e., a Dynamic Convolutional Block (DCB), two cascaded Wavelet transforms, Enhancement Blocks (WEBs) and a Residual Block (RB). | Improved results compared to some popular denoising methods |
| Darooei et, al.(14) | 2022 | Segmentation | OCT | Dual Tree Complex Wavelet (DTCW) | Application of DTCW sub bands as the input of a U-net. | 1) More robust<br>2) An improved time-frequency analysis of data that has been successfully applied to deep learning segmentation tasks |
| Wang et, al.(15) | 2022 | Classification | Natural images (Caltech-256) | DTCW & WPT | Application of a CNN model with wavelet domain inputs | Improvement of the network's classification performance significantly |
| Sarhan et, al.(16) | 2020 | Classification | MRI | Wavelet (Haar) | Extraction of features from images by utilizing the strong energy compactness property exhibited by the Discrete Wavelet Transform and application in a CNN | High reduction of the dimensions of the input image simplifies the work of the CNN classifier |
| Lakshmanaprabu et, al.(17) | 2019 | Classification | CT images | Discrete Wavelet Transform (DWT) | Utilizing wavelet features along with histogram and texture features. Then, using the reduced features by Linear Discriminant Analysis (LDA) as the input of Optimal Deep Neural Network (ODNN) which is optimized with Modified Gravitational Search Algorithm (MGSA). | Time saving |
| Mohsen et, al.(18) | 2018 | Classification | MRI | DWT (Haar) | Application of DWT to extract the features of the segmented tumors and use them as Deep Neural Network (DNN) input | 1) Requires fewer hardware specifications<br>2) Takes a convenient time for processing large-size images |
| Khatami et, al.(19) | 2017 | Classification | Radiography images | 2D-DWT (Haar) | Application of 2D-DWT to capture the highly discriminative coefficients that represent the complex structure of original data. Then, using the best selected coefficients by Kolmogorov Smirnov (KS) Test as the input of Deep Belief Network (DBN) feature extractor | 1) Time-consuming reduction<br>2) Increasing Accuracy |
| Rezaeilouyeh et, al.(20) | 2016 | Classification | histopathology images of breast & prostate tissues | shearlet | Extraction of the magnitude and phase of shearlet coefficients and feeding these extra features along with the original images to the CNN | Application of shearlet transform as a general mathematical tool and extracting features without any hand-crafting |
| Williams et, al.(21) | 2016 | Classification | Natural images (MNIST & CIFAR-10) | Wavelet | Application of preprocessed data in the wavelet domain as the input of CNN | Substantial increase in accuracy compared to the spatial domain processing |

In the current study, we will investigate the suitability of different X-let transforms in CNN for the classification of AMD, DME, and Normal OCT B-scans. To find the most efficient X-let transform, first, all the B-scans are transformed to the different sparse multi-scale X-let domains and all the sub-bands have been fed to a CNN model in parallel to extract and select the best features. These features then have been utilized as the input of a Multi-Layer Perceptron (MLP) and a Support Vector Machine (SVM) individually as classifiers.

The rest of this paper is arranged as follows: Section 2 describes the utilized datasets, X-let transforms, proposed CNN, and classifiers. Section 3, and 4 present experimental results and gives some discussions, respectively. Our work is concluded in Section 5.

## 2. Materials and Methods

### 2.1 Database

#### 2.1.1 Heidelberg Dataset

The first dataset (Dataset-A) used in this study was acquired and collected by Heidelberg SD-OCT imaging systems at Noor Eye Hospital in Tehran(22). This dataset consists of 50 normal (1535 B-scans), 48 AMD (1590 B-scans), and 50 DME (1054 B-scans) subjects. The data information is summarized in the supplementary, Table 1.

#### 2.1.2 Basel Dataset

The second dataset (Dataset-B) utilized in this research was collected in Didavaran eye clinic, Isfahan, Iran using an SS-OCT imaging system designed and built in Dep. of Biomedical Engineering, University of Basel(23). According to the classification application that is the main goal of our investigation, the “Aligned-Dataset QA” was chosen which has been obtained after image contrast enhancement, denoising, and alignment of raw data, respectively(23). In this study, 17 DME (2338 B-scans), 15 Non-diabetic (2492 B-scans), and 19 normal (2169 B-sacns) cases were selected manually among 40, 50, and 34 available subjects, respectively, because their B-scans are almost clear and suitable for classification application.

### 2.2 Data Preprocessing

According to Rasti et al. (22) which provides a proper preprocessing algorithm for Dataset-A, the following steps were applied in this study, with the difference that here the investigations are based on a B-scan not a volume. We added a denoising step to this preprocessing algorithm since it was shown experimentally that this leads to better results. Figure 2 (a) displays the different steps of the preprocessing algorithm.

**Figure. 2.**
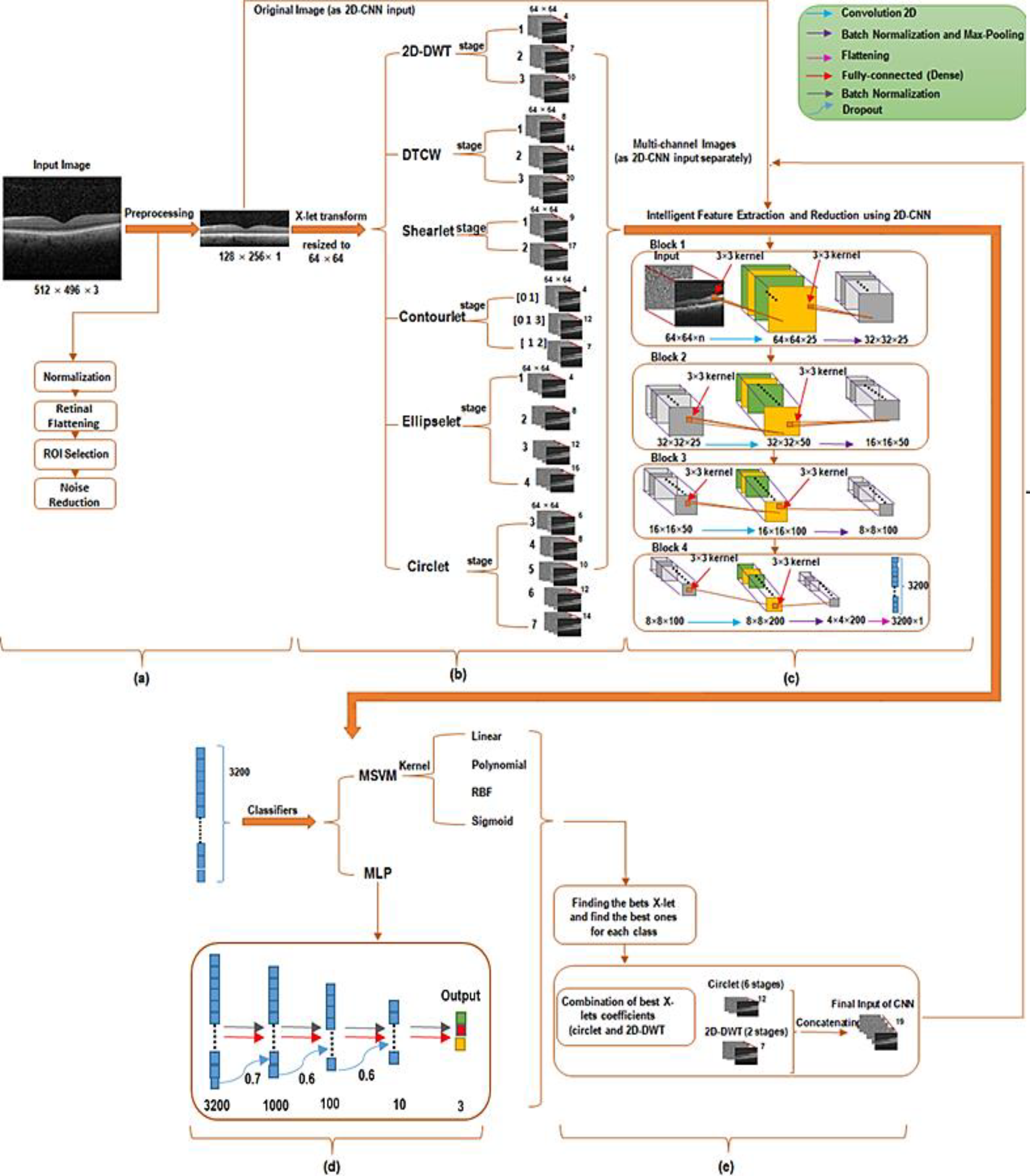
The framework of the proposed classification methods. Where parts (a) to (d) indicate the steps of preprocessing, transformation, feature extraction, and classification respectively.

1. Normalization: First, all B-scans were resized to 496×512×1 pixels to make the field of view of OCT images unique. Then, data normalization was applied by dividing of each B-scan into 255, such that all pixel intensities are in [0, 1].
2. Retinal Flattening: Next, a curvature correction algorithm (35) was used, where the hyper-reflective-complex (HRC) is detected as the whole retinal profile, and then localized using the graph-based geometry.
3. Region of Interest (ROI) Selection: Then each B-scan was cropped vertically by selecting 200 pixels above and 100 pixels below the detected HRC. These values were chosen manually in order to focus on the region of the retina containing the main morphological structures while preserving all retinal information. Next, cropped B-scans were resized to 128 × 512 pixels and the ROI for each one was selected by cropping a centered 128 × 470 pixel subimage. Finally, each selected ROI was resized to 128 × 256 pixels for further processes.
4. Noise Reduction: Noise Reduction: Ultimately, denoising of the data was achieved using a non-local means algorithm with a deciding filter strength of 10.

For the “Aligned-Dataset QA” that was employed as Dataset-B, some steps of preprocessing algorithm such as Retinal Flattening and Noise Reduction were already done. The size of each B-scan was 300 × 300 pixels. Therefore, each B-scan was divided by 255, then it was cropped horizontally by omitting the first 50 pixels (since they have no remarkable information) and finally resized to 128 × 256 pixels.

Example B-scans of each class for both datasets, before and after preprocessing, are presented in the supplementary file Figure 1.

### 2.3 Splitting Training-set and Test-set

Any correlation between test and train images can cause bias and affect the results. Hence, to avoid this unwanted leakage, all the images belonging to one subject should be considered as only the test-data or training-data. Ultimately, test data and train data were divided using 5-fold nested-cross-validation.

### 2.4 Classification Strategy

#### 2.4.1 X-let Transforms

The main purpose of this study is to compare the effect of different geometrical X-let transforms, in two or higher dimensions, for OCT classification. These transforms are provided by directional time-frequency dictionaries(24), and they give powerful insight into an image’s spatial and frequency characteristics. X-lets are available mathematical tools that provide an intuitive framework for the representation and storage of multi-scale images(21).

Therefore, in this study several geometrical X-let transforms including 2D-DWT (11, 25, 26) (Note that Haar wavelet was taken in current study), DTCWT (14, 27) (Note that just the real parts of this transform are utilized in this research to reduce the complexity and redundancy), shearlets (28, 29), contourlets (30), circlets (31), and ellipselets (24) were applied to decompose each B-scan a linear combination of basis functions or dictionary atoms. The non-subsampled (NS) (32) form of the multi-scale X-let transforms was employed to build a multi-channel matrix for each B-scan using all the sub-bands in parallel. Each multi-channel matrix was ultimately resized to (64×64×number of channels) pixels in order to reduce computational complexity and save time. The details of all utilized X-lets are summarized in the supplementary file, Table 2.

**Table. 2.**
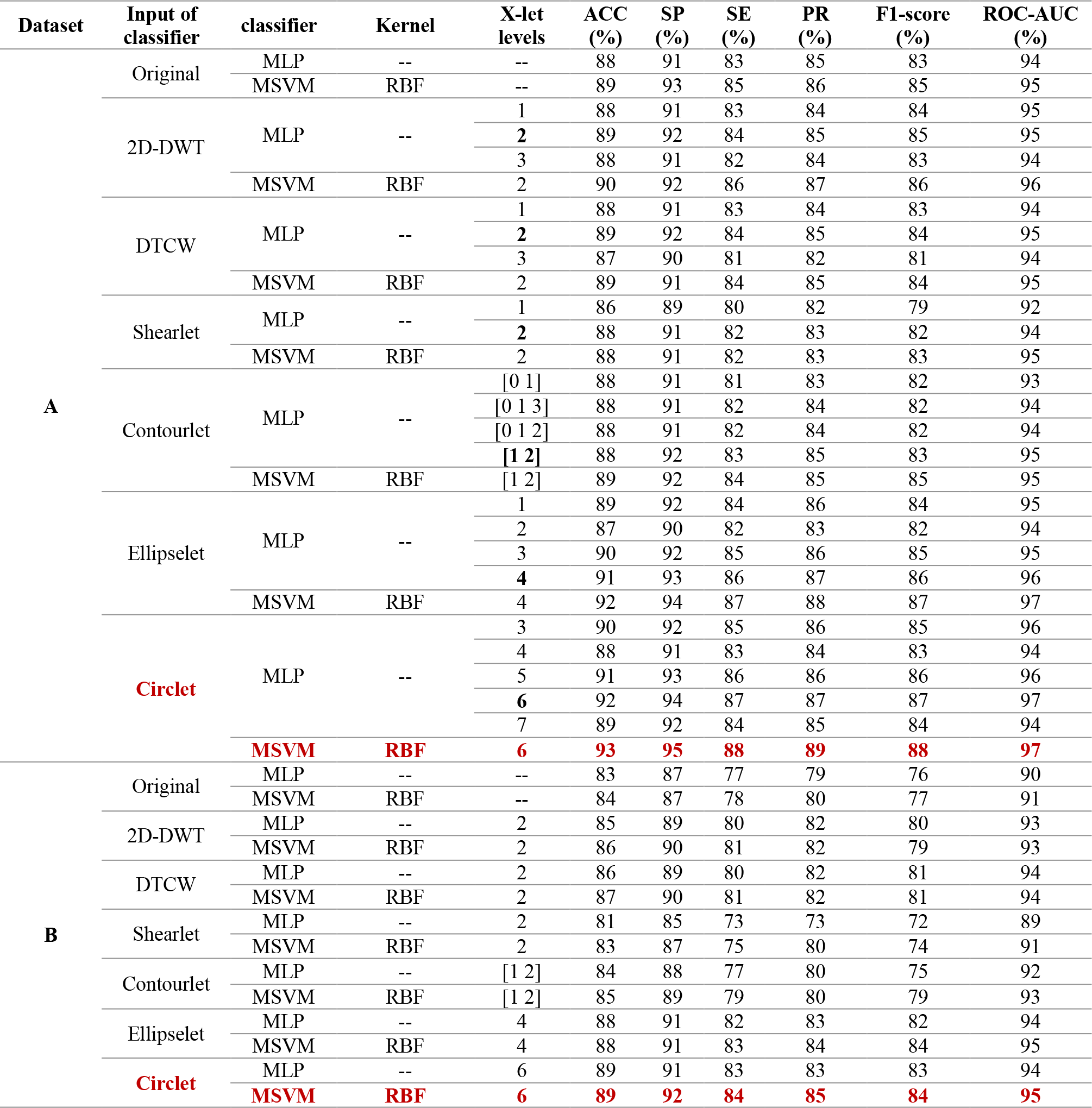
Evaluation criteria for different numbers of levels for each X-let transform using MLP classifier and the combination of the best number of X-let levels and RBF as the best kernel of MSVM classifier. Black bold values show the best number of levels for each X-let while red bold values indicate the best values, the best classifier, and the best X-let for OCT classification.

#### 2.4.2 Intelligent Feature Extraction

Using a large amount of features for a large number of training-set as the input of NN-models can cause high computational complexity. Therefore, using a proper feature reduction algorithm can reduce the training time, improve the accuracy by decreasing the redundant data and thereby reduce the over-fitting (33).

According to the 2D nature of the data and, it is essential to employ using an algorithm that is able to extract 2D features is essential. Most neural networks and deep learning algorithms convert the 2D input into a vector of neurons. In contrast, 2D-CNNs are optimized for 2D pattern recognition problems and pay attention to the 2D nature properties of images (34), which makes them best suited for image classification (21, 35). 2D-CNN itself has the ability to extract all the features of OCT images in all directions but using the X-let transformed datas as its input can control the feature extraction effectively and make it more intelligent.

Therefore, we designed a 2D-CNN where the last convolutional layer was flattened and employed as final features which were fed into the classifiers.

As shown in the Figure 2 (b) and (c), multi-channel images obtained using each X-let transform were used as the proposed CNN input. The architecture of this CNN (as shown in Figure 2 (c)) consists of 4 blocks including the 2D-convolution-layer (CL), Batch-Normalization (BN), and Maximum-pooling layers. The filter size of each CL was set to 25, 50, 100, and 200 for blocks 1 to 4, respectively, while “ReLU” activation function (AF), zero padding, and the kernel size of 3×3 were utilized in each CL. The optimizer and the loss function were tuned to “Adam” and “categorical-cross-entropy”, respectively. The hyper-parameters in this model were tuned manually on learning-rate of 10^-3^, batch-size of 8, and maximum-epoch of 100, with the aim of achieving the highest possible accuracy in the training phase.

#### 2.4.3 Classifiers

Next, a Multi-Class Support Vector Machine (MSVM) and a Multi-Layer Perceptron (MLP) were applied as classifiers separately. MSVM was used because it is known as a simple classifier that can help to investigate the importance and the direct effect of each X-let transform in results. On the other hand, deep-learning-based architectures have been successfully applied recently in the field of biomedical image processing (9, 36-38). Since a 2D-CNN was used for feature extraction, subsequently an MLP algorithm for OCT B-scans classification can be used in order to apply a fully deep-learning method.

##### MSVM

SVM is a simple classifier and can give some perspective on the performance of different X-lets, that predicts two classes by finding a hyper-plane that best separates the two classes (39, 40). When the data is perfectly linearly separable, Linear SVM is suitable. Otherwise, kernel tricks can help for classification. A kernel function (such as a radial-basis-function (RBF), polynomial (poly), or sigmoid) tries to convert the lower dimension space (which is not linearly separable) to a higher dimension, where a decision boundary can be found more easily. Although, the SVM technique inherently was developed to classify just two classes, it can be upgraded to a multi-class classifier using some techniques. One of these techniques is known as the One Versus One (OVO) strategy, which was applied in this study. The OVO strategy divides the dataset into one dataset for each class versus every other class as shown in the supplementary file, Figure 2. Ultimately, a voting system, determines which class is accurate for each B-scan (41).

The Grid Search algorithm was used to find the best hyper-parameters for each kernel. This algorithm calculates the accuracy of each combination of hyper-parameters in each kernel and selects the values which provide the best accuracy. These tuned values are summarized in the supplementary file, Table 3.

**Table. 3.** Performance comparison of the proposed CNN and VGG19 and VGG16 as feature extraction models. These values were obtained using a combination of Circlet and 2D-DWT bases as the input to the models. Where Red Bold values represent the best-achieved ones for each dataset.

| Feature Extraction model | Dataset | ACC (%) | SE (%) | SP (%) | PR (%) | F1-score (%) | ROC-AUC (%) |
| --- | --- | --- | --- | --- | --- | --- | --- |
| **Proposed CNN** | A | **94.5** | **96** | **89.5** | **90** | **90** | **98** |
|  | B | **90** | **92** | **84.5** | **86** | **85** | **96** |
| VGG19 | A | 93 | 94.5 | 88 | 88 | 88 | 96.5 |
|  | B | 88.5 | 91 | 83 | 84 | 84.5 | 95 |
| VGG16 | A | 91.5 | 93 | 87.5 | 87 | 87 | 96 |
|  | B | 87 | 90 | 82 | 83 | 82.5 | 94 |

##### MLP

MLP is a nonlinear multi-layer feed-forward neural network that follows a supervised learning technique known as the backpropagation learning algorithm (42). In this study, the output of CNN was fed into MLP as the input layer. Three hidden layers (fully connected (FC)-BN) with 1000, 100, and 10 neurons, respectively, were used. With the purpose of reducing over-fitting probability, an optimized dropout factor of 70%, 60%, and 60% were considered for the hidden layers, respectively. For hidden layers, “ReLU” AF was employed, however, for the output layer, an FC layer with 3 neurons and “Softmax” AF was utilized. The optimizer and loss function and hyper-parameters were selected similarly to those in the proposed CNN. Figure 2 (d) indicates the MLP classifier architecture.

### 2.5 Classification Evaluation

K-fold cross-validation is a powerful means for testing the performance of machine learning models (43). A reliable accuracy estimation will have a relatively small variance across folds (44). However, one drawback of this method is that the split in each fold is done completely randomly. To address this problem, Stratified-K-fold cross-validation can be employed in which instead of a random split, the division is done in the way that the ratio between the target classes in each fold is the same as in the full dataset (45, 46). In the current study, a nested form of Stratified-K-fold was used in order to split test, validate and train data in each fold. We have chosen K=5, therefore, the experiment was conducted 5 times, the evaluation parameters were calculated in each fold on the test dataset, and the average values of all folds were reported as the final results.

The following evaluation parameters including accuracy (ACC), sensitivity (SE), specificity (SP), precision (PR), F_1_-score, and area-under-the-Receiver-Operating-Characteristic (ROC) curve (known as ROC-AUC) were calculated for each X-let transform.

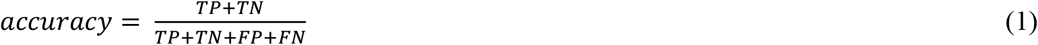

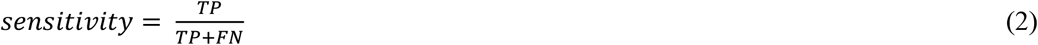

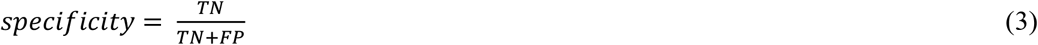

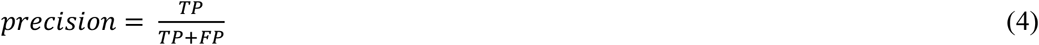

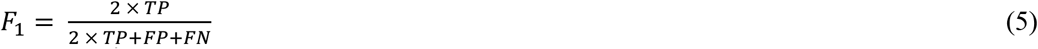

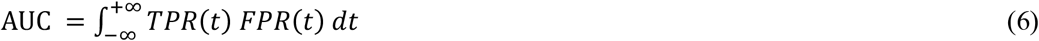

Here TP is the true positive, FN is the false negative, TN is the true negative, and FP is the false positive. TPR and FPR define the true positive rate and the false positive rate respectively.

## 3. Experimental Results

In the current study, MATLAB R2020a software was used to extract contourlet, circlet, and ellipselet representations and the Keras and Tensorflow platform backend in python 3.7 software environment was employed to extract 2D-DWT, DTCW, and shearlet coefficients. Classification models were also implemented in this environment.

In order to find the best kernel of MSVM, the precision-recall curves for all classes using different kernels were plotted in Figure 3, where the original Dataset-A was considered as input of MSVM. The P-R curve shows the tradeoff between precision and recall for different thresholds. The average of the area under these curves for all classes is written below the curves in each subplot. A high P-R-AUC represents both high precision and high recall which are related to a low false negative rate and a low false positive rate, respectively. As shown in Figure 3, the average P-R-AUC of classes for sigmoid, polynomial, and linear is equal to 0.89, 0.9, and 0.9, respectively, while it is equal to 0.92 for RBF kernel which indicates the better performance of this kernel in the classification of this dataset.

**Figure. 3.**
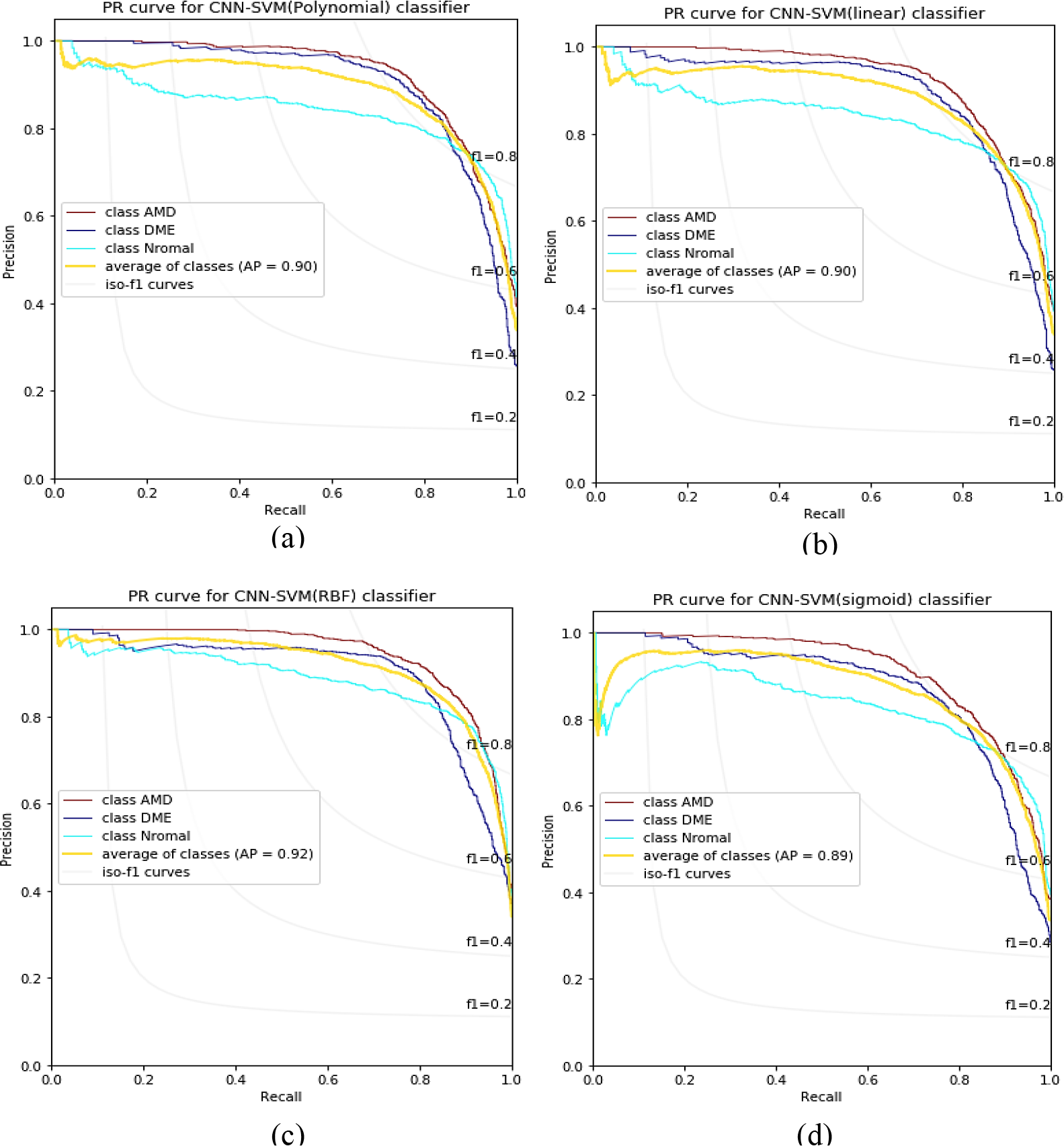
P-R curves of classes for each kernel of MSVM using original Dataset-A. (a) to (d) represent the polynomial, linear, RBF, and sigmoid kernel respectively.

As the next step, we examined the performance of different stages of each X-let transform for the classification of Dataset-A using the MLP. Considering RBF as the best kernel of MSVM, evaluation parameters were also reported using the RBF-MSVM and the best number of stages for X-lets. This step was repeated using the proposed MLP and RBF classifier and the best number of X-let levels for Dataset-B. Table 2 shows these results. As mentioned in Section 2.4.1, for the contourlet transform, the decomposition level were shown in a vector.

## 4. Discussion

According to Table 2, it is evident that the circlet transform performs better than the other X-let transforms. The best confusion matrices related to RBF-MSVM classifier and circlet transform for both datasets are shown in the supplementary file, Figure 3.

The accuracy achieved for each X-let transform in classification of each class individually, is presented in Figure 4, where the circlet transform achieves the best performance for the DME class while the 2D-DWT provides a better result for Normal class. It seems that the appearing circles on DME B-scans can be detected much better using the circlet transform (Figure 5 shows some of these appearing circles caused by the accumulation of fluid). Moreover, Figure 6 shows the ROC curves of classes for the circlet transform and the 2D-DWT. We observe that the ROC-AUC of the DME class is better using circlet transform whereas the Normal class has a better ROC-AUC using the 2D-DWT because most of the B-scan layers belonging to this class are aligned at 0 degrees.

**Figure. 4.**
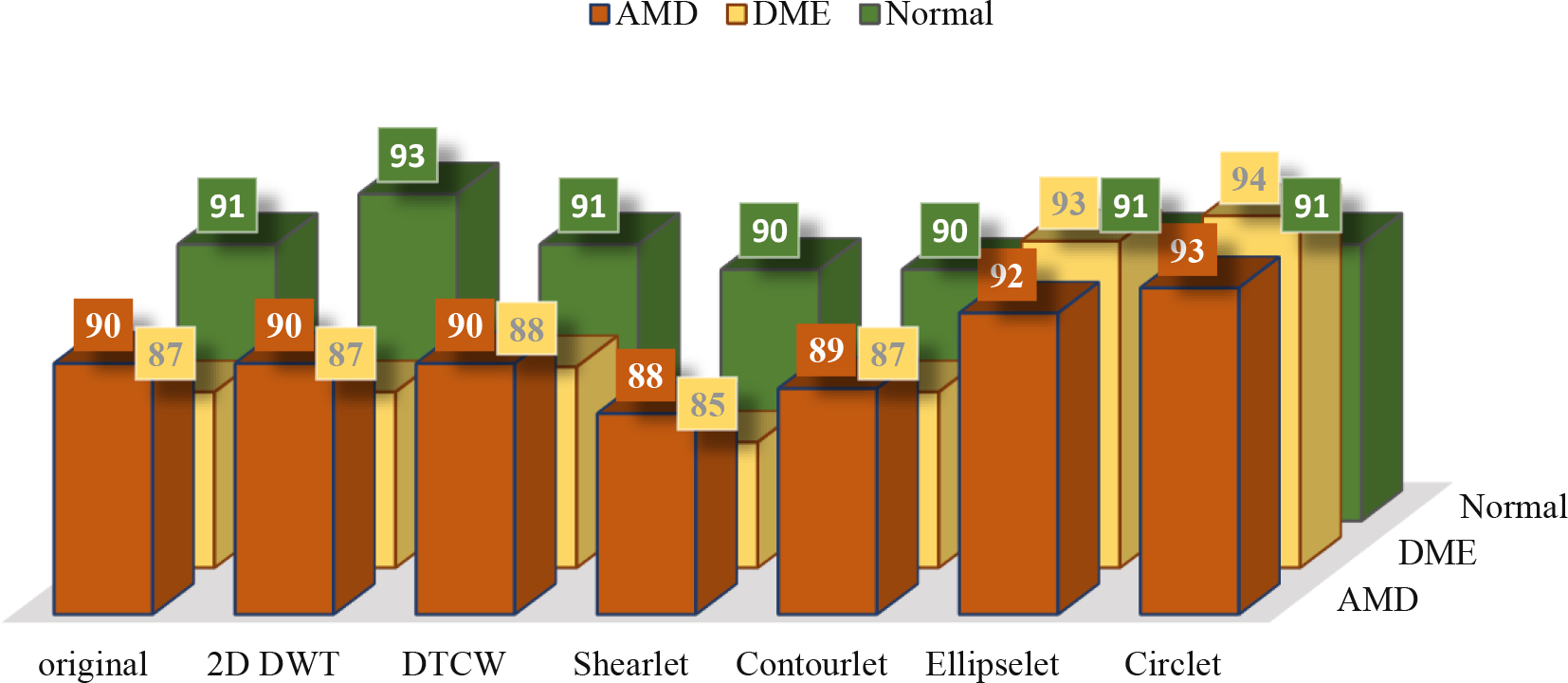
The accuracy of DME, AMD, and normal classes using MSVM with RBF kernel and different X-lets. These values are expressed as a percentage.

**Figure. 5.**
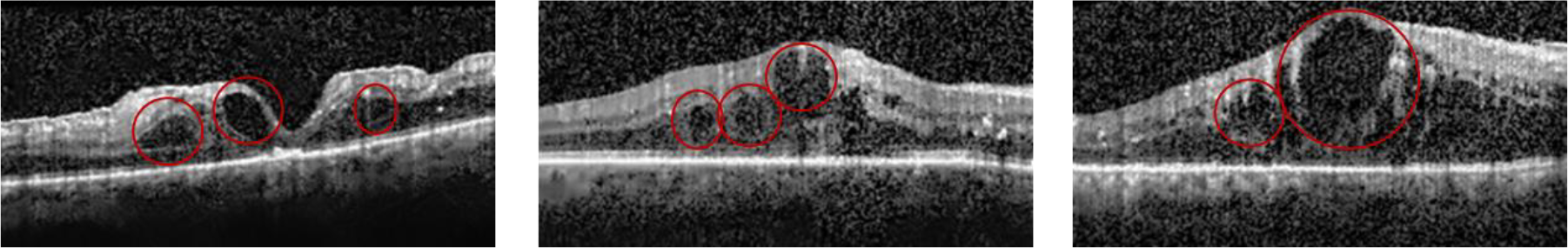
Appearing circles on B-scans of DME subjects. Because of the fluid accumulation.

**Figure. 6.**
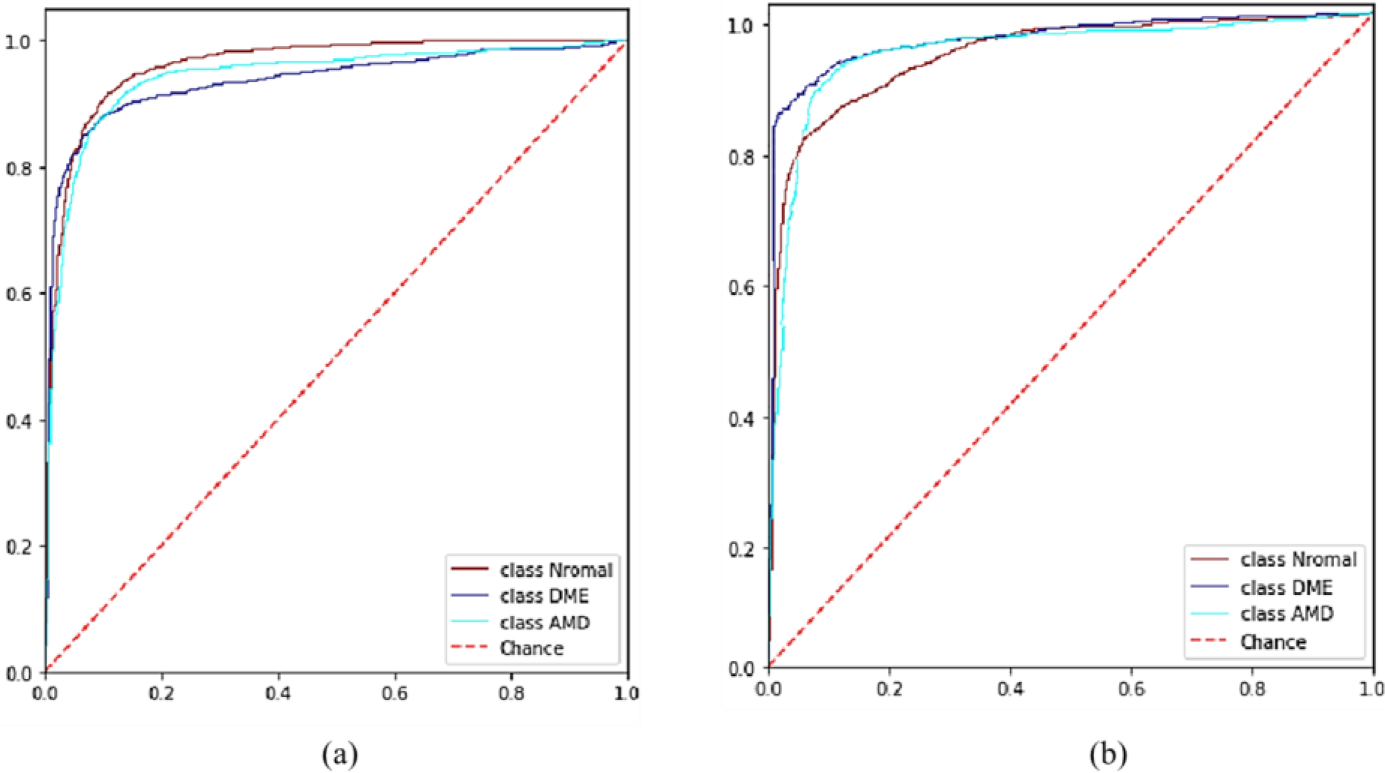
The ROC curves of different classes for (a) 2D-DWT, and (b) circlet transform. Where the ROC curve of normal, DME, and AMD classes are shown in dark red, purple, and turquoise color respectively.

In order to compare the classification results achieved by employing the two transforms (2D_DWT, and circlet) with classification using the original image, the B-scans were reconstructed using half of the sub-bands of each transform individually. The reconstructed B-scans were again utilized as the input of proposed models. Finally, the Grad-CAM class activation visualization was plotted in Figure 7 for several B-scans using the original B-scans and the reconstructed ones using circlet transform bases (for DME cases) and the 2D-DWT (for Normal cases), respectively. This heat map can give some perspective of the parts of an image with most impact for the classification score. For the reconstructed B-scans using the circlet transform these image parts concentrate on appearing circles on DME B-scans, for the reconstructed B-scans using 2D-DWT, these image parts contain lines in Normal B-scans, while these image parts in original B-scans do not focus on these characteristics. Note that for all B-scans shown in Figure 7, the classifier predicted the class correctly using either the original data or the X-let transforms, but the heat maps concentrations are totally different.

**Figure. 7.**
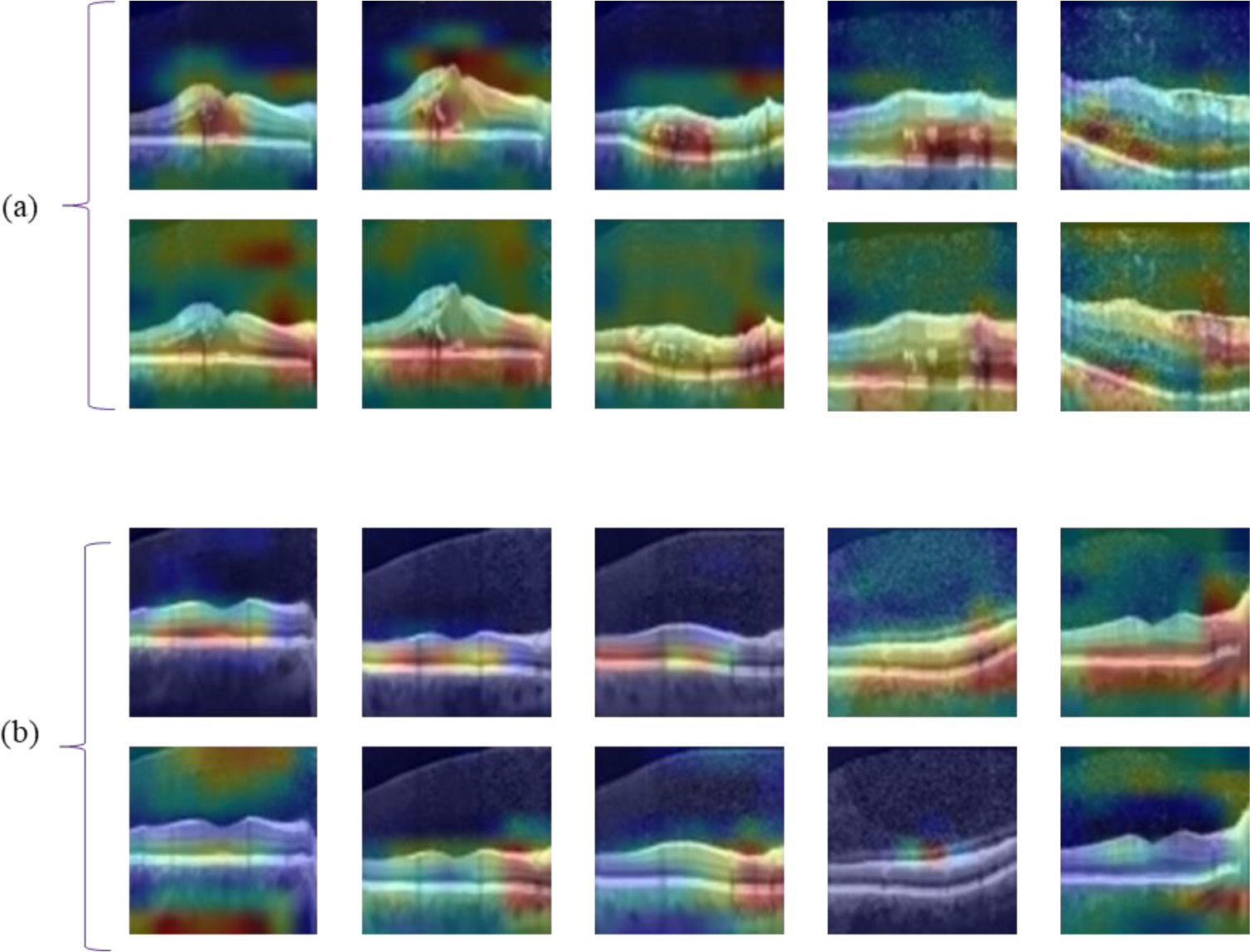
The Grad-Cam of the proposed CNN for several test data of Dataset-A. Where part (a) compares the heat maps of reconstructed B-scans using circlet bases (first row) and the associated original B-scans (second row) and part (b) compares the heat map of reconstructed B-scans using 2D-DWT bases (first row) and the associated original B-scans (second row). Note that part (a) shows DME B-scans and part (b) shows normal B-scans.

Accordingly, it seems that a combination of these two transforms can provide a better performance in the classification of these datasets because it can simultaneously emphasize the nature of circles in the DME class and straight lines in the Normal class. Results of this experiment are reported for both datasets in Table 3. In addition, in this table the performance of the proposed CNN is compared to VGG16 and VGG19 as two state-of-the-art models. Note that, these values are obtained using MSVM with RBF kernels as the best proposed classifier.

According to Table 3, the proposed CNN performs better than VGG19 and VGG16 in feature extraction (although it uses much fewer trainable parameters). Furthermore, the combination of the circlet transform and 2D-DWT provides better results than using only the circlet transform.

We call our proposed transform and method the “CircWave” and the “CircWaveNet”, respectively. To demonstrate the advantages of CircWave compared to the Circlet and 2D-DWT, three B-scans from each class were selected and the heat map was plotted in Figure 8 for each one, where the reconstructed B-scans use half of the sub-bands of each of the three mentioned transforms, separately, were utilized as the CNN input. That is clearly obvious that for the Normal case the 2D-DWT provides a more accurately focused heat map, while for the DME case, the Circlet performs better. However, the proposed CircWave transform can concentrate on the correct regions for both Normal and DME cases simultaneously, and provides a more suitable heat map for AMD cases as well. Note that these three B-scans were classified correctly using all three mentioned transforms.

**Figure. 8.**
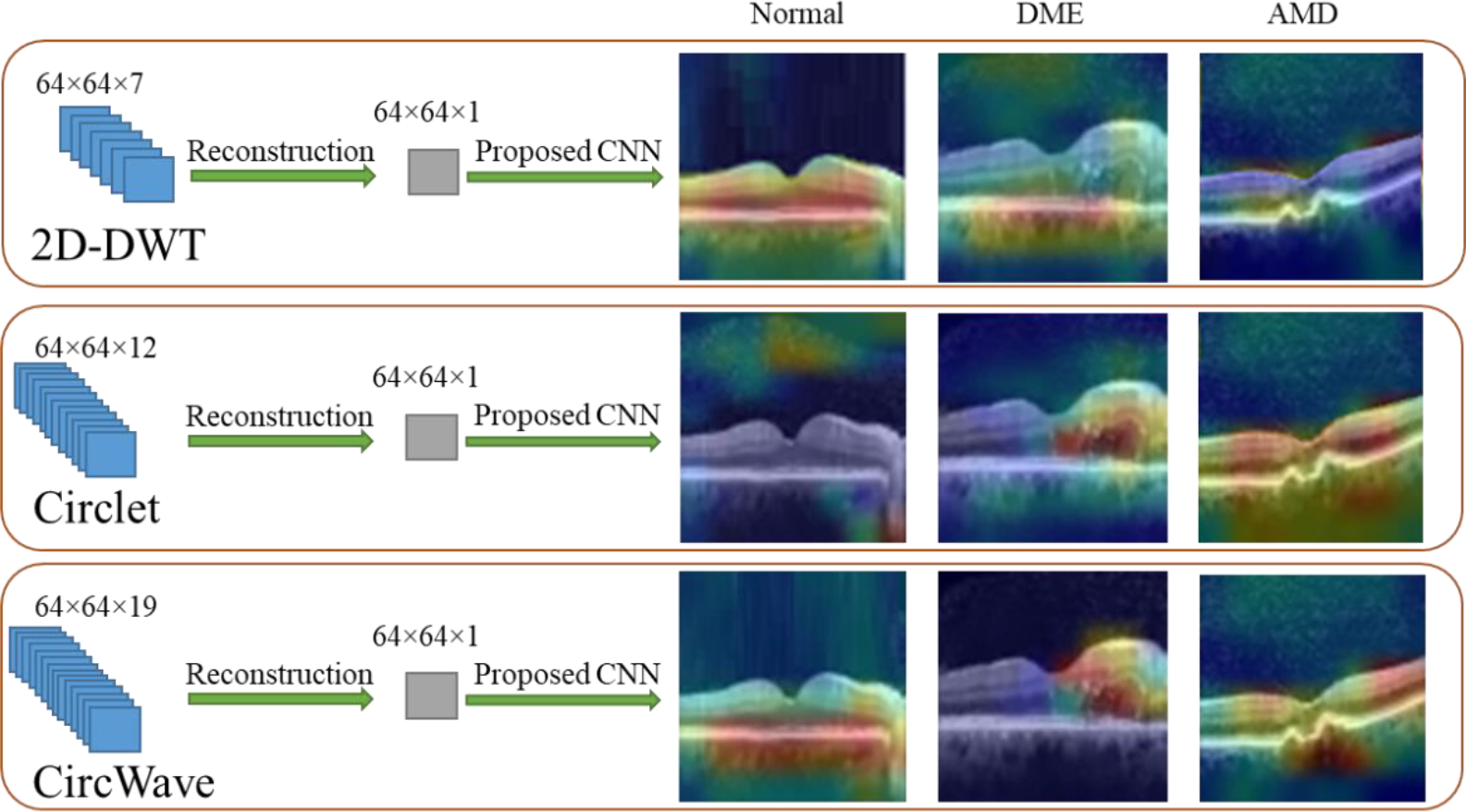
The Grad-Cam of the proposed CNN for three test data of Dataset-A of each class, when reconstructed B-scans using 2D-DWT, Circlet, and CircWave transforms (in first to third rows), respectively are used as the input of the CNN.

We also used Principal Component Analysis (PCA) plus T-distributed stochastic neighbor embedding (t-SNE) techniques to visualize the high-dimensional outputs of the proposed CNN, when original data, 2D-DWT transformed data, Circlet transformed data, and CircWave transformed data were used as inputs, respectively. The proposed CNN has 3200 dimensions output for each mentioned input. First, the PCA reduction algorithm was employed to reduce the number of dimensions and created a new dataset containing the fifty dimensions. Then, they were reduced again to two dimensions using the t-SNE technique. Finally, the dataset created from each input was plotted in Figure 9. It can be noticed that when CircWave bases are used as CNN input, samples of all three classes are spaced apart and well grouped together with their respective cases. While the Circlet transform can cluster DME cases very clearly in their own class, it cannot space apart Normal and AMD cases well. Also, the 2D-DWT has the ability to group Normal cases well but cannot separate AMD and DME cases properly. Obviously, the original data is not efficient in providing separable features for this 3-class classification problem.

**Figure. 9.**
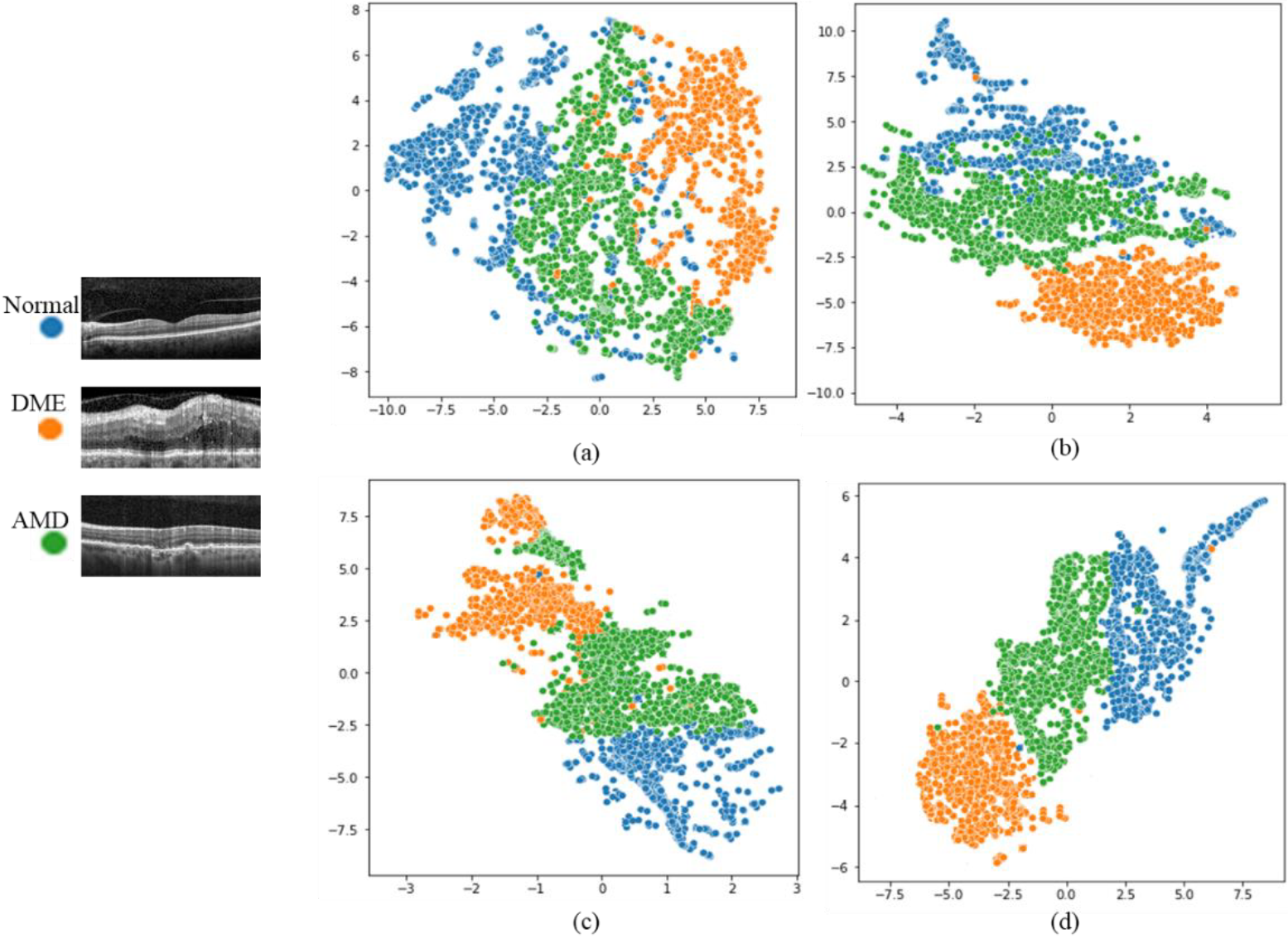
Visualization of the output of the proposed CNN using PCA plus t-SNE reduction algorithms for (a) The original data, (b) the Circlet bases, (c) the 2D-DWT bases, and (d) the CircWave bases.

To the best of our knowledge, it was the first time that the Dataset-B was utilized for the classification application and we have shown that the proposed CircWaveNet is successful in classification of this dataset and outperforms the results obtained using the original data and other transforms.

On the other hand, there are several research results that use the Dataset-A for classification application. In Table 4, we summarize these results.

**Table. 4.** Results of other articles on Dataset-A. Results of the proposed CircWaveNet are shown in bold.

| Number | Paper | Dimension | Method | ACC (%) | SE (%) | PR (%) | ROC-AUC (%) |
| --- | --- | --- | --- | --- | --- | --- | --- |
| 1 | Rasti et, al.(22) | 3D | Multi-scale Convolutional Mixture of Expert | -- | -- | 99.39 | 99.8 |
| 2 | Fang et, al.(47) | 3D | Lesion-Aware CNN | -- | 99.36 | 99.39 | 99.80 |
| 3 | Das et, al(48) | 3D | B-scan Attentive CNN | 93.2 | -- | -- | 95 |
| 4 | Rasti et, at(49) | 3D | Wavelet-based Convolutional Mixture of Experts | -- | -- | -- | 99.3 |
| 5 | Wang e, al(50) | 3D | Volumetric OCT-Recurrent Neural Network | 93.8 | 94.0 | 94.4 | -- |
| 6 | Wang et, al.(50) | 2D | CliqueNet | 98.6 | -- | -- | -- |
| 7 | Das et, al.(51) | 2D | semi-supervised<br>Generative Adversarial Network | 97.43 | 97.43 | -- | -- |
| 8 | Xu et, al(52) | 2D | Multi-branch Hybrid Attention Network | 99.7 | -- | 1 | -- |
| 9 | Nabijiang et, al(53) | 2D | Block Attention Mechanism | 99.64 | -- | -- | -- |
| **10** | **Out Method** | **2D** | **CircWaveNet** | **94.5** | **96** | **90** | **98** |

According to the Table 4, some of the results seem superior to those of CircWaveNet, but one should note that the first four mentioned articles worked on 3-D volumes. In their method, they use a specific threshold (like: τ =15 or τ =30) and if more than τ percentages of B-scans belonging to one subject are predicted as abnormal, the maximum probability of B-scans’ votes (according to AMD or DME likelihood scores) determines the type of patient retinal disease. The fifth article used volume-level labels for each subject (instead of labeling each B-scan separately). By contrast, in this paper, the parameters are achieved based on the B-scan which is much more difficult than a subject-based one.

The other articles (no. 6 to 9) mentioned in Table 4, as well as this paper, worked on 2D B-scan classification. Although these papers obtained better results than our method, it should be noted that in these articles, the train and test sets are divided according to the ratio of 8:2, regardless of the possible leakage between test and train subjects, which causes bias and certainly increases the results that are classified wrongly. However, in this article, all the images belonging to one subject were considered as only the test-data or training-data. This data splitting strategy makes the test results more reliable for ophthalmologists.

Besides, most of the mentioned methods need to train a large amount of training parameters (more than 100 million) while in CircWaveNet the total training parameters are approximately 3.5 million which greatly reduces the computational complexity.

## 5. Conclusion

In this paper, we proposed to apply suitable X-let transforms of OCT B-scans rather than the original images as input for the 2D-CNN to achieve improved classification results while strongly reducing computational cost. This is possible by transferring the data to a transform domain that allows a sparse image representation with a small amount of transform coefficients. Ice have shown that almost all X-let transforms can lead to more accurate classification results than the original B-scans. Among all utilized X-let transforms, the circlet transform performs better for both considered datasets obtaining 93% ACC, 95% SE, 88% SP, 89% PR, 88% F1-score, and 97% ROC-AUC in Dataset-A and 89% ACC, 84% SE, 92% SP, 85% PR, 84% F1-score and 95% ROC-AUC in Dataset-B. Concentrating on class-accuracy, we found that the 2D-DWT can perform better for the classification of Normal cases because most lines and boundaries in a normal B-scan almost follows a straight pattern with zero degrees which can be well detected using simple 2D wavelet transform that is able to extract lines with 0, 90 and ±45 degrees. However, in the retinal structure of DME cases, some circles appear due to fluid accumulation and an increase in retinal thickness. This characteristic changes the pattern of B-scans of DME cases that is extracted much better using the circlet transform. Moreover, it has been shown in this paper that these two transformations not only provide a significant increase of evaluation parameters but also focus on the characteristics of each class that are very important for ophthalmologists. Since it is necessary but not sufficient for them to categorize each case, X-lets make this decision more reliable because they concentrate on the true discriminative features of each class. Despite the classifier can predict the class for most B-scans using even original data, the CNN based on the considered X-let transforms focuses exactly on the features that make a difference in classes, while for original data this is not the case.

As the next step and in order to increase the accuracy of classification models, 2D-DWT and circlet transform coefficients were concatenated and fed to the models. This proposed algorithm increased the evaluation parameters by about 0.5 to 1.5 percent in both datasets.

It has been shown in this paper that despite extensive progress of deep learning in image classification, there are limitations in some cases, including medical images, due to the lack of data or labeled data. Therefore, image processing techniques such as the application of time-frequency transforms can help to make this application more accurate and reliable since it can analyze and interpret the data and make decisions based on the truth.

In future research, data-adaptive transform models can be employed to match the non-data-adaptive ones (X-lets) to classifiers as much as possible and extract adapted dictionaries for sparse image representation and feature selection for classification application. It is expected that data adaptive transforms that are fed with the best non-data adaptive transforms, increase the accuracy of classifiers.

## Data Availability

All utilized data are available online at:
https://misp.mui.ac.ir/en/dataset-oct-classification-50-normal-48-amd-50-dme-0
and
OCT Basel Data | Medical Image and Signal Processing Research Center

https://misp.mui.ac.ir/en/oct-basel-data-0

