## Supplementary for "CircWaveNet: A New Conventional Neural Network Based on Combination of Circlets and Wavelets for Macular OCT Classification"

This supplementary provides detailed information about the data, the proposed method, and the experimental results.

Table.1. Heidelberg dataset information

| **Characteristics** | **Value** | **Unit** |
|---|---|---|
| Axial resolution | 3.5 | μm |
| Scan-dimension | 8.9×7.4 | $mm^2$ |
| Number of A-scans | 512 to 768 | Scans |
| Number of B-scans per volume (for different patients) | 19, 25, 31, and 61 | Scans |

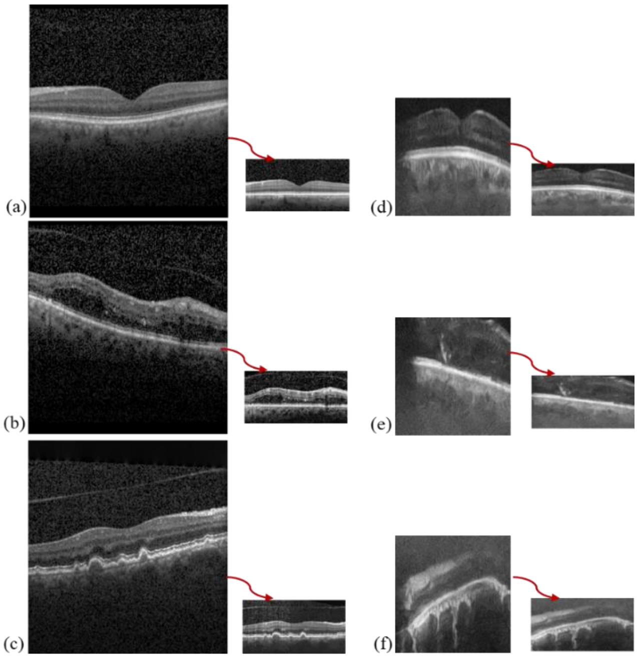

Figure. 1. Example B-scans of (a) Normal, (b) DME, and (c) AMD cases in Dataset-A, and (d) Normal, (e) Diabetes, and (f) non-Diabetes in Dataset-B, before and after preprocessing. Red arrows connect the non-preprocessed B-scans to the related preprocessed ones.

Table. 2. Properties of Different X-lets

<table>
<tr><th>X-let</th><th colspan="2">Properties</th></tr>
<tr><td rowspan="2">2D-DWT (NS)</td><td>Note</td><td>First applying a one level Haar wavelet to each row and secondly to each column</td></tr>
<tr><td>Filter Bank (1 stage)</td><td>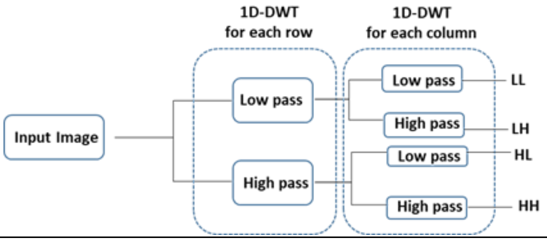
</td></tr>
<tr><td rowspan="2">DTCW (NS)</td><td>Note</td><td>1. Using two real DWTs in parallel to create real and imaginary parts of the transform.<br>2. Producing six sub-bands in six directions (±15, ±45, and ±75)</td></tr>
<tr><td>Filter Bank (1 stage)</td><td>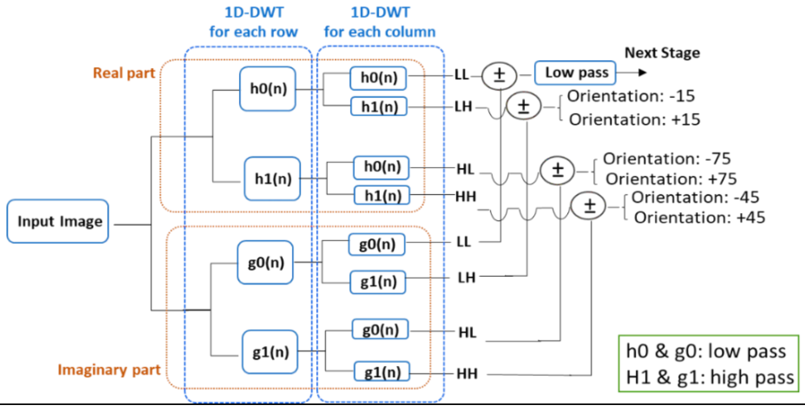
</td></tr>
<tr><td rowspan="3">Contourlet (NS)</td><td>Note</td><td>1. Using multi-directional decomposition and multi-scale decomposition for images.<br>2. Divided into two shift-invariant parts: a Non Subsampled Pyramid structure (that ensures the multiscale property) and a Non Subsampled Directional Filter Bank structure (that gives directionality).</td></tr>
<tr><td>Filter Bank</td><td>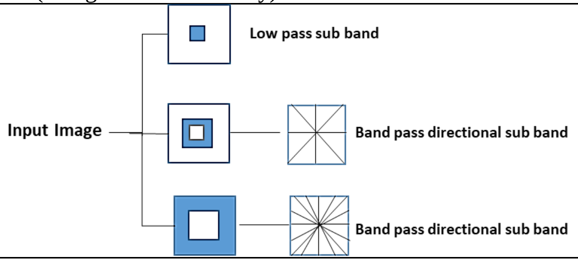
</td></tr>
<tr><td>Idealized frequency partitioning</td><td>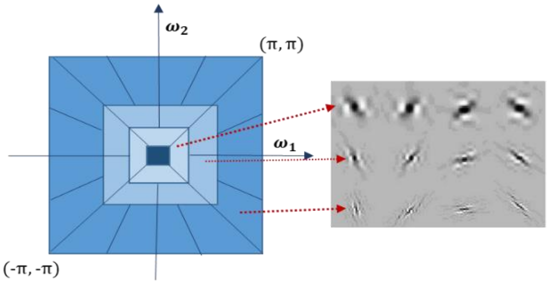
</td></tr>
<tr><td>Shearlet (NS)</td><td>Note</td><td>1. Similar to Contourlet but instead of directional filters, shearlet filters are used.<br>2. Steps: (a) Multi-scale decomposition (achieved by NSP), (b) Localization of orientation (accomplished by a shearlet Filter)</td></tr>
</table>

| | | |
|---|---|---|
| | Filter Bank | 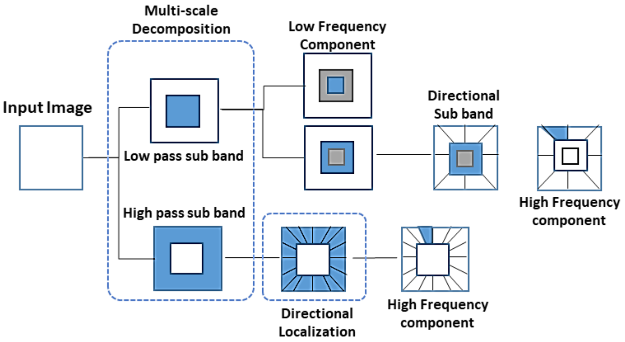<br> |
| **Circlet** | Note | 1. A robust tool to detect circular objects in images without the need for image segmentation<br>2. Decomposes an image to a set of circles with different radii using a Discrete Fourier Transform (DFT) filter bank<br>3. Describing the circlet parameters by a central position (x0, y0), radius (r0), and central frequency content (f0)<br>4. Obtaining all circlet components (Cμ(x, y)) by a shift or change in radius and central frequency in a reference circlet (Cref(x, y)) |
| | Basis Functions | 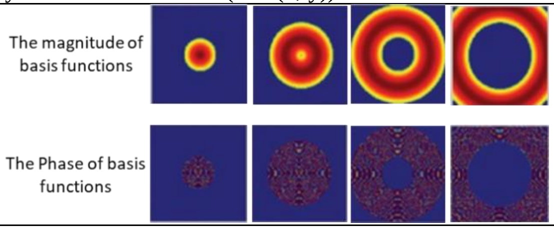<br> |
| | Idealized frequency partitioning | 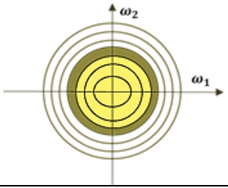<br> |
| **Ellipselet** | Note | 1. An extension of the circle transformation with elliptic basis functions<br>2. With four basis functions in four different directions at angels 0°, 90°, −45°, and 45° |
| | Basis functions | 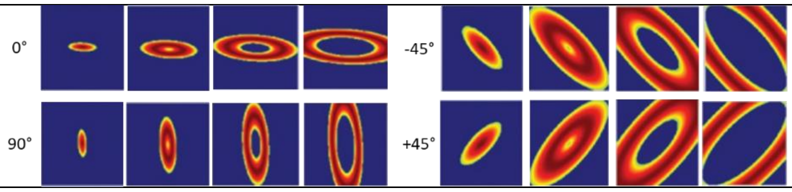<br> |
| | Idealized frequency partitioning | 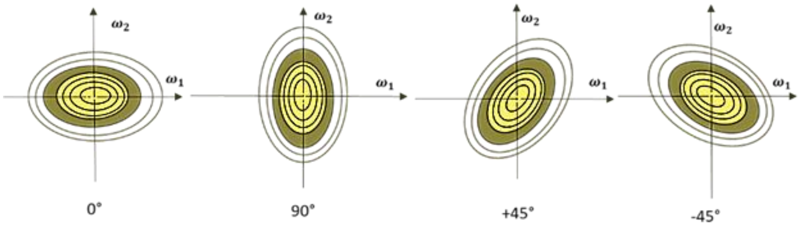<br> |

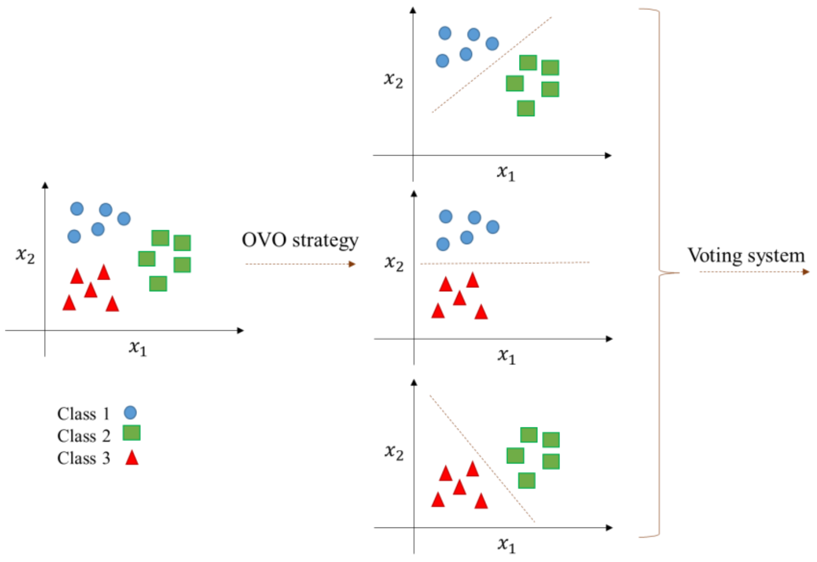


Figure. 2. OVO strategy in MSVM

Table.3. Hyper-parameters selected for different kernels by Grid Search algorithm

| kernel | Hyper-parameter | Tuned value |
|---|---|---|
| RBF | C | 100 |
| | gamma | $10^{-7}$ |
| Linear | C | 0.01 |
| Polynomial | C | 10 |
| | Degree | 2 |
| Sigmoid | C | 10 |

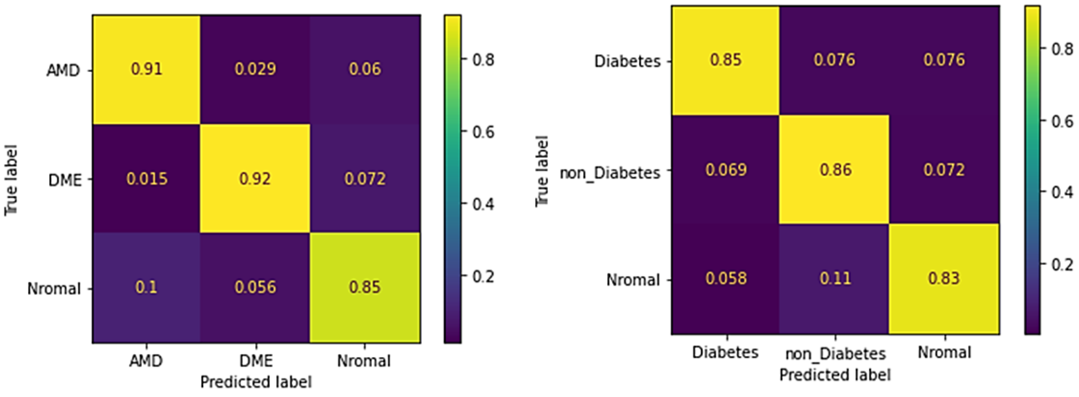


(a) (b)

Figure. 3. Confusion matrix of MSVM classifier with RBF kernel for (a) Dataset-A and (b) Dataset-B, while the input of the classifier is the circlet basis functions of B-scans.
